# The discriminative ability of ROTEM for Delayed Cerebral Ischemia and Poor Clinical Outcome following Aneurysmal Subarachnoid Haemorrhage

**DOI:** 10.1101/2023.09.11.23295389

**Authors:** Maud A. Tjerkstra, Homeyra Labib, Bert A. Coert, René Post, W. Peter Vandertop, Dagmar Verbaan, Nicole P. Juffermans

## Abstract

**Introduction:** Aneurysmal subarachnoid haemorrhage (aSAH) and delayed cerebral ischemia (DCI) have been associated with hypercoagulability as detected by viscoelastic testing. In this study, we evaluate temporal alterations in rotational thromboelastography (ROTEM-) coagulation profiles and the discriminative ability of ROTEM-parameters for DCI and poor clinical outcome following aSAH.

**Materials and Methods:** ROTEM-parameters were measured on days 0, 3-5 and 9-11 after aSAH and compared between patients with and without DCI, radiological DCI and 6-months’ poor clinical outcome (modified Rankin Scale 4-6). ROC-curve analyses were used to calculate areas-under-the-curve (AUC) and to determine optimal cut-off values with a sensitivity of >90% and highest possible specificity for DCI and radiological DCI. For poor outcome, a specificity >90% with highest possible sensitivity was used.

**Results:** Of 160 aSAH patients, 31 (19%) had DCI, 16 (10%) radiological DCI and 68 (44%) had poor outcome at six months. DCI, radiological DCI and poor clinical outcome were associated with hypercoagulability. The ROTEM-parameter with the best discriminative ability for radiological DCI was INTEM CT (AUC: 0.75), with optimal cut-off value <153 seconds (sensitivity 94%, specificity 59%). For poor outcome, this was FIBTEM A10, (AUC: 0.85), with optimal cut-off value >27 mm (specificity 94%, sensitivity 49%).

**Conclusions:** Hypercoagulability, as detected by ROTEM-parameters, is an excellent marker of poor clinical outcome after aSAH and might be useful for stratifying patients for inclusion in future trials on therapeutic interventions. Conversely, the absence of hypercoagulability on ROTEM may be used to identify patients at low risk of DCI for early hospital discharge.

## INTRODUCTION

Aneurysmal subarachnoid haemorrhage (aSAH) is a devastating disease with a case fatality of 32-42% and a permanent disability rate of approximately 50% amongst survivors.^1^ In patients who survive the first 24 hours, the most important contributor to poor outcome is delayed cerebral ischemia (DCI), occurring in approximately one out of three to four aSAH patients, mostly between days three to 12 post-SAH.^2–5^ In order to monitor for DCI, patients with aSAH are generally hospitalized for at least 14 days. The pathophysiology of DCI is incompletely understood, but thought to be multifactorial, including the formation of microthrombi.^6–9^ A reliable diagnostic test for DCI is still lacking.^10,11^ Clinically, DCI is diagnosed by exclusion, or based on radiological criteria, which becomes apparent only days after onset of neurological deterioration.

The involvement of microthrombosis suggests a hypercoagulable state in patients who develop DCI compared to those who do not. Thus far, individual markers of coagulation and fibrinolysis have shown no predictive association with DCI.^12^ Rotational thromboelastometry (ROTEM) analyses whole blood and provides information about the contribution of coagulation factors, fibrin formation, platelet function and fibrinolysis.^13^ Studies have shown a hypercoagulable ROTEM-profile in aSAH patients compared to controls, already present early after ictus and worsening over time.^14–19^ To date, two studies have evaluated ROTEM in DCI patients and showed hypercoagulability within 72 hours following SAH.^14,18,20^ These studies evaluated group differences of a few ROTEM-parameters only, and were limited by small sample sizes, hampering determination of cut-off values.

The aim of this study is to evaluate the discriminative ability of ROTEM in patients with aSAH by determining specific cut-off values for DCI and poor clinical outcome at six months, defined as a modified Rankin Scale (mRS) score of 4-6.

## METHODS

### Study design

We performed a prospective cohort study from October 2018 to May 2023 at the Amsterdam UMC. The study was interrupted during the COVID-pandemic. Amsterdam UMC is a tertiary referral centre for aSAH patients in the Amsterdam metropolitan area. Consecutive patients were included during daytime if they met the following criteria: 1) patients aged 18 or older, 2) diagnosis of a non-traumatic SAH by either subarachnoid blood on non-contrast head CT or a positive lumbar punction, and if 3) blood collection for this study was possible within 24 hours after ictus and before aneurysm treatment. For this study, we excluded patients with non-aneurysmal SAH. The study was conducted in accordance with the Declaration of Helsinki. The protocol was approved by our Institutional Review Board (MEC 2017_318). Deferred informed consent was obtained from all patients, or their legal representatives, in exception from those who suffered imminent death. Standard of care comprised of securing the aneurysm < 24-72 hours by endovascular coiling if technically feasible, or by surgical clipping. All patients received 2850 IE fraxiparin as thrombosis prophylaxis. During the first part of the inclusion period, a randomized controlled trial on ultra-early and short tranexamic acid treatment was conducted in the same patient group.^21^

### Data collection

We used the prospectively collected data of the institution’s SAH-registry, which comprised the entire hospitalization period (at least 14 days) and follow-up clinical outcome assessment at six months after SAH. The following variables were used in this study: baseline characteristics (including the World Federation of Neurological Surgeons grade and Fisher grade); common complications following aSAH, and use of anti-haemostatic agents during hospitalization; and clinical outcome assessed by the mRS at six months after aSAH, which was scored by a trained nurse using a structured and validated interview at the outpatient clinic or by telephone. Additional laboratory values on admission, including hemoglobin, CRP and leukocytes, were retrospectively collected from the electronic patient records.

### Blood sampling

Citrated BD Vacutainer® tubes were collected as soon as possible after symptoms of SAH (day 0; T0) and on days 3-5 after aSAH (T1). For the first 98 patients, we also collected blood on days 9-11 after aSAH (T2), if possible. Blood was used for ROTEM-analysis within two hours after withdrawal.

### Coagulation tests

Using ROTEM® Sigma (Werfen, Benelux), a citrated blood sample was inserted into a cartridge containing INTEM, EXTEM, and FIBTEM assays. The INTEM assay contained ellagic acid as activator of the intrinsic pathway, the EXTEM assay contained tissue factor as activator of the extrinsic pathway, the FIBTEM assay contained tissue factor and cytochalasin D to block platelet contribution and assess fibrinogen contribution to clot formation. Of the EXTEM and INTEM assays we used: clotting time (CT), clot formation time (CFT), α-angle, amplitude at 10 minutes (A10), maximum clot firmness (MCF) and clot lysis index at 60 minutes (LI60). Of FIBTEM assays we used α-angle, A10 and MCF. CT depicts the time until clot initiation, which reflects coagulation factor function. CFT and α-angle depict the speed of clot formation, i.e. clot kinetics, reflecting activity of intrinsic clotting factors, fibrinogen cross-linking and platelet function. A10 and MCF depict the clot strength at 10 minutes after CT and the maximum clot strength, respectively, reflecting the fibrinogen and platelet function (Figure 1).

**Figure 1.**
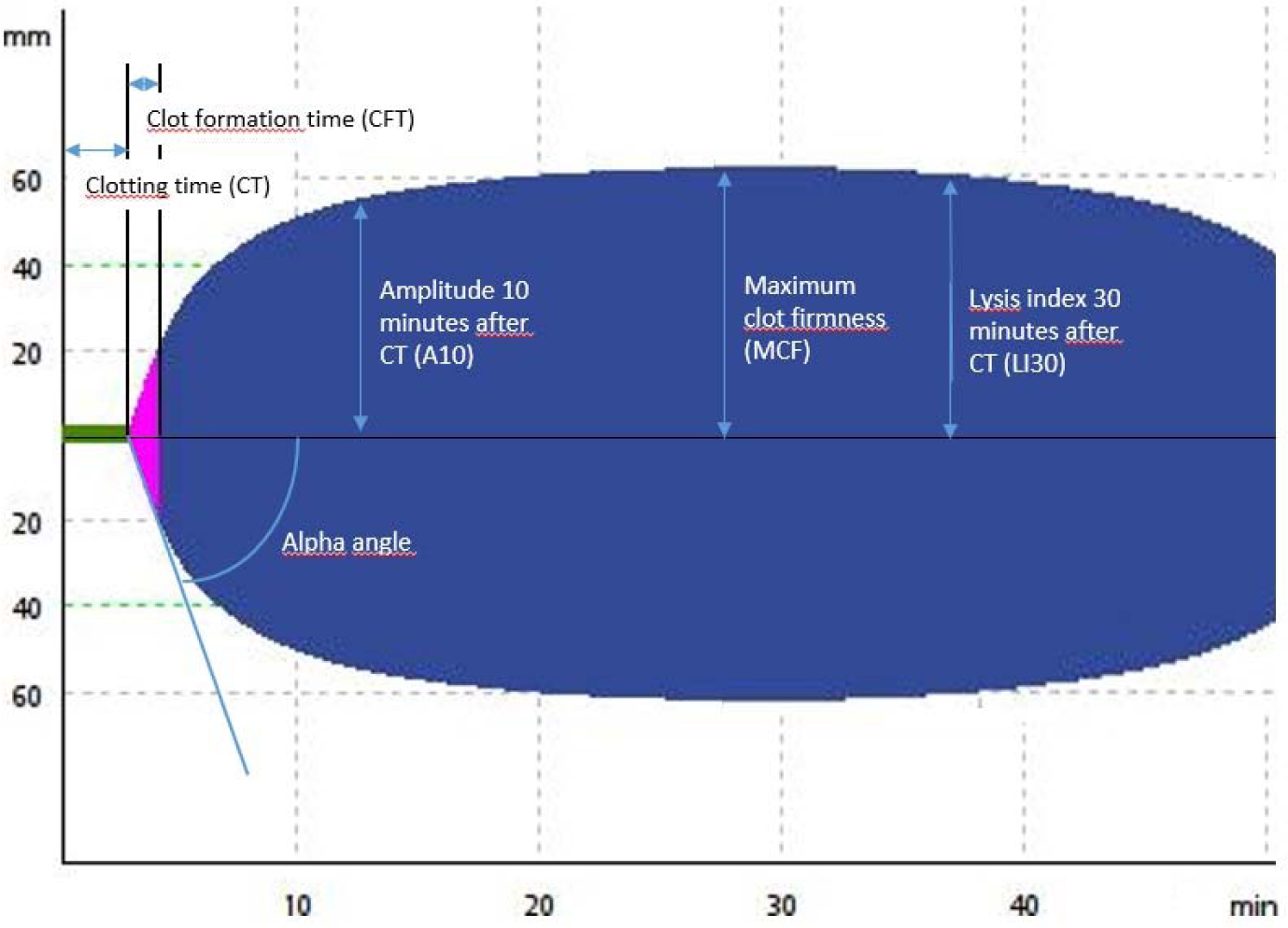
Depiction of ROTEM-parameters

Conventional coagulation parameters were measured with Sysmex CS-2500 and included PT, aPTT, fibrinogen and D-dimer. The stored citrate plasma samples were thawed serially in a 37 degrees Celsius water bath for 10 minutes and vortexed to mix the plasma thoroughly. The coagulation parameters were measured as soon as possible. The following reagents were used: Innovin PT reagents (Siemens B4212-100), Actin FS (Siemens B4218-100) and CaCl2 25mMol/L (Siemens ORHO375) for aPTT, Thrombin Reagent (Siemens B4233-25) for fibrinogen and Innovance D-Dimer (Siemens OPBO07). The detection limits were 7.0 to 180.0 seconds for PT, 20.0 to 180.0 seconds for aPTT, 0.3 to 8.9 g/L for fibrinogen and 0.2 to 35.2 mg/L for D-dimer. If a measurement exceeded the detection limit, the value was set at the detection limit.

### Outcome measurements

The primary outcome was DCI, defined as either clinical or radiological DCI, or a combination of both, according to the definitions proposed by Vergouwen et al. in 2010.^22^ In short, the definition for clinical DCI was: “the occurrence of a decrease in consciousness or focal neurological impairment, which lasts for at least one hour and cannot be attributed to any other cause” and for radiological DCI: “the presence of cerebral infarction on CT-or MR-imaging, which was not present on CT-or MR-imaging within 24-48 hours after SAH and cannot be attributed to the aneurysm treatment or other causes”. CT-or MR-imaging was only performed on indication.

As the clinical diagnosis of DCI is merely a description of symptoms and therefore, especially in complex SAH cases with a multitude of complications, prone to misinterpretation, we used radiological DCI (irrespective of neurological symptom onset) as secondary outcome measurement. Also, poor clinical outcome (mRS 4-6) at six months was used as secondary outcome.

### Definitions

Rebleeding was scored when patients suddenly deteriorated with increase in blood pressure suggestive for rebleeding or in the presence of an increase of SAH on CT-imaging compared to a previous investigation. Treatment complications were categorized into haemorrhagic, thrombo-embolic and ischemic complications. Haemorrhagic complications were defined as either extravasation of contrast dye or perforation of the micro catheter, -wire or coil during endovascular treatment or active bleeding during neurosurgical treatment. Thrombo-embolic complications were defined as reduced passage or stasis of contrast in an artery or slowed venous outflow without the aspect of vascular spasm and was evaluated by the treating neurointerventional radiologist. Ischemic complications were defined as post-procedural neurological deterioration compared to the pre-operative neurological status, in combination with new ischemic lesions on post-operative imaging. Hydrocephalus was defined as a gradual onset of deterioration of consciousness with CT evidence of either enlarged ventricles or increased intracranial pressure measured by a lumbar puncture. Seizures were scored positively if patients had a clinical event suggestive for seizures (at the discretion of the treating physician) and were subsequently administered anti-epileptic drugs. Meningitis was defined as a positive cerebrospinal fluid culture. Pneumonia was defined as either a positive sputum culture and/or an infiltrate on chest x-ray. Urinary tract infection was defined as a positive urine culture or abnormal urine sample.

### Statistical analyses

Baseline characteristics were provided as means with standard deviation (SD), medians with interquartile range (Q1-Q3) or proportions (%), depending on the type and distribution of data. Normality of continuous variables was tested by the Shapiro-Wilk test (threshold for normality; W>0.9). To evaluate the temporal alterations in coagulation profiles, we assessed group differences in ROTEM- and conventional coagulation parameters on all three time points for patients with and without DCI, using either the independent T-test or the Mann-Whitney U test. Statistical significance was set at p<0.05. Additionally, a sensitivity analysis, comparing the value of ROTEM-parameters in groups after exclusion of patients who received tranexamic acid in the context of the ULTRA-trial,^21^ was done. Missing data was handled using a pairwise deletion method.

Parameters with significant differences in the primary group comparison (including all patients) were selected for calculation of odds ratios (OR) with corresponding 95% confidence intervals (95% C.I.) by univariate logistic regression. For the logistic regression analyses, we included only ROTEM measurements which had been obtained prior to DCI-onset. As the median onset of DCI is generally five to seven days after aSAH, we decided to not include ROTEM measurements obtained on day 9-11. ROTEM-parameters which were significantly associated with DCI in univariate logistic regression analyses were used to calculate the area-under-the-curve (AUC) with corresponding 95% C.I. In general, an AUC of 0.5 suggests no discriminative ability, an AUC 0.7-0.8 is considered acceptable, 0.8-0.9 excellent and >0.9 outstanding.^23^ The ROTEM-parameter with the best discriminative ability was defined as the ROTEM-parameter with the highest AUC. For this ROTEM-parameter, the sensitivity and specificity were calculated for all points on the curve to determine an optimal cut-off value.^24–26^ For DCI, it would be of interest to identify patients who will not develop DCI, for whom hence earlier hospital discharge would be a safe option. In this instance, a false negative test result could result in early discharge of a patient who will develop DCI. A test with high sensitivity will have a high proportion of true positives, may also have a substantial proportion of false positives, however, it would hardly ever be false negative. Since our patient cohort is not very large, a sensitivity threshold of 100%, or >95%, seems not purposeful, as the accompanying specificity will likely be quite low and the determined cut-off value will have a poor positive predictive value and very low number of negative tests, which will hinder early discharge of patients.^27^ Therefore, the optimal cut-off value was determined by the point on the curve with a value for sensitivity of at least 90%, and subsequent highest possible specificity. If more than one ROTEM-parameter had the highest AUC, the ROTEM-parameter with the best discriminative ability was considered the ROTEM-parameter with the highest combination of sensitivity and specificity.

Similar analyses were performed for radiological DCI and poor clinical outcome, however for poor outcome we included measurements of all three time points and used different criteria to determine the optimal cut-off value. For clinical outcome, it would be of interest to accurately predict poor outcome in an early stage of the disease, thereby (in combination with other clinical, radiological or biochemical factors) potentially supporting physicians in decisions on determining futility of care. In this instance, a false positive (poor outcome) test result could support physicians in the decision to forego life-sustaining treatments, whereas the patient would have had a good clinical outcome. A test with high specificity for poor clinical outcome, will have a high proportion of true negatives, may also have a substantial proportion of false negatives, however, it would hardly ever be false positive. Therefore, for poor clinical outcome, an optimal cut-off value was determined by the point on the curve with a value for specificity of at least 90% and subsequent highest possible specificity.

## RESULTS

The cohort comprised of 160 aSAH patients with a mean age of 57.8 years (SD 12.5), of whom 114 (71%) were female, 79 (50%) presented with WFNS grade 4-5 and 156 (97%) patients had Fisher grade 3-4. DCI occurred in 31 patients (19%) at a median of nine days (range 2 - 16) after aSAH. DCI-onset was prior to the second blood collection in 4/30 (13%) DCI patients (excluded for logistic regression and ROC analyses). Radiological DCI was seen in 16 (10%) patients (prior to second blood collection in 1/16) and 68 (44%) patients had poor outcome. ROTEM-measurements were available for 157/160 patients (98%) on admission, 125/160 (78%) patients on days 3-5 and 63/160 (39%) on day 10 after aSAH. Reasons for missing ROTEM-measurements are listed in Figure 2. Twenty-six aSAH patients received ultra-early and short-term (<24h) tranexamic acid as participant of the ULTRA-trial. None of the baseline characteristics and common complications following aSAH, except from treatment modality (p=0.03), significantly differed between patients with and without DCI (Table 1).

**Figure 2:**
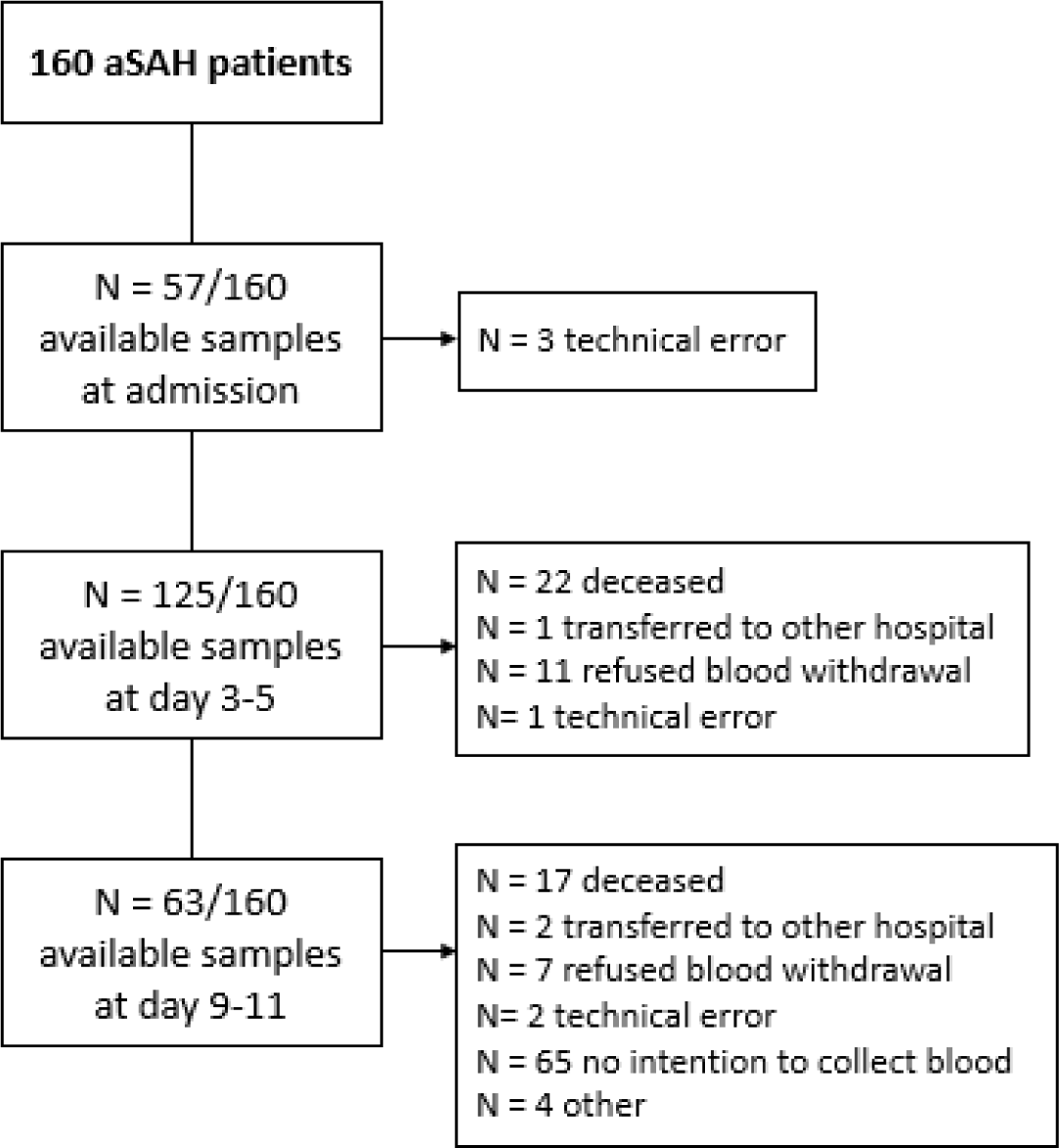
Available blood samples on admission, days 3-5 and days 9-11 after ictus of 160 aSAH patients.

**Table 1:** Baseline characteristics and common complications following aSAH of 160 patients with aSAH.

|  | aSAH (n=160) | DCI (n=31) | No DCI (n=129) | P-value |
| --- | --- | --- | --- | --- |
| **Age** mean (SD) | 57.8 (12.5) | 58.0 (13.0) | 57.8 (12.4) | 0.92 |
| **Female** | 114 (71) | 27 (87) | 87 (67) | 0.05 |
| **Comorbidities\*** |  |  |  |  |
| Diabetes mellitus | 5 (3) | 2 (7) | 3 (2) | 0.24 |
| Hypertension | 48 (30) | 13 (43) | 35 (28) | 0.12 |
| Hypercholesterolemia | 10 (6) | 2 (7) | 8 (6) | 1.00 |
| Cardiovascular disease | 14 (9) | 1 (3) | 13 (10) | 0.31 |
| **Smoking status<sup>†</sup>** |  |  |  | 0.21 |
| Current | 56 (51) | 13 (12) | 43 (49) |  |
| Previous | 25 (23) | 7 (6) | 18 (21) |  |
| Never | 30 (27) | 3 (3) | 27 (31) |  |
| **Home medication** |  |  |  |  |
| Anti-platelet | 15 (9) | 4 (3) | 11 (7) | 0.49 |
| Anti-coagulation | 5 (3) | 0 (0) | 5 (4) | 0.58 |
| **Tranexamic acid administration**<br>(ULTRA-trial intervention group) | 26 (16) | 4 (13) | 22 (17) | 0.78 |
| **WFNS** |  |  |  | 0.76 |
| Grade 1 | 50 (31) | 8 (26) | 42 (33) |  |
| Grade 2 | 26 (16) | 4 (13) | 22 (17) |  |
| Grade 3 | 5 (3) | 1 (3) | 4 (3) |  |
| Grade 4 | 25 (16) | 7 (23) | 18 (14) |  |
| Grade 5 | 54 (34) | 11 (36) | 43 (33) |  |
| **Fisher** |  |  |  | 0.56 |
| Grade 2 | 4 (3) | 0 (0) | 4 (3) |  |
| Grade 3 | 31 (19) | 7 (23) | 24 (15) |  |
| Grade 4 | 125 (78) | 24 (77) | 101 (78) |  |
| **Treatment modality** |  |  |  | 0.03 |
| Endovascular | 124 (78) | 29 (94) | 95 (74) |  |
| Surgical | 14 (9) | 2 (7) | 12 (9) |  |
| None | 22 (14) | 0 (0) | 22 (17) |  |
| **Admission laboratory<sup>‡</sup>** |  |  |  |  |
| Hemoglobin (mmol/L, mean (SD)) | 8.3 (1.04) | 8.1 (1.1) | 8.3 (1.0) | 0.52 |
| Leukocytes, ($\cdot 10^9/L$, mean (SD)) | 12.3 (5.1) | 14.0 (6.4) | 12.9 (4.6) | 0.06 |
| CRP (mg/L, median (IQR)) | 2.0 (0.9-4.6) | 2.6 (1.0-5.4) | 1.9 (0.8-4.1) | 0.25 |
| **Complications** |  |  |  |  |
| Rebleeding | 30 (19) | 7 (23) | 23 (18) | 0.61 |
| Hydrocephalus | 113 (71) | 25 (81) | 88 (68) | 0.20 |
| Treatment-related |  |  |  |  |
| Haemorrhagic complications | 6 (4) | 3 (10) | 3 (3) | 0.13 |
| Thrombo-embolic complication | 6 (4) | 1 (3) | 4 (4) | 1.00 |
| Cerebral ischemia | 5 (4) | 0 (0) | 6 (6) | 0.34 |
| Meningitis | 11 (7) | 4 (13) | 7 (5) | 0.23 |
| Pneumonia | 21 (13) | 3 (10) | 18 (14) | 0.77 |
| Urinary tract infection | 10 (6) | 1 (1) | 9 (7) | 0.69 |
| Seizures | 22 (14) | 4 (13) | 18 (14) | 1.00 |

aSAH: aneurysmal subarachnoid haemorrhage, DCI: delayed cerebral ischemia, WFNS: World Federation of Neurological Surgeons. \* n=3 (2%) missing, <sup>†</sup> n=49 (31%) missing, <sup>‡</sup> hemoglobin: n=14 (9%) missing, leukocytes: n=13 (8%) missing, CRP: n=55 (34%) missing

### DCI

Except from EXTEM CT, all ROTEM-parameters and conventional coagulation markers, at all time points, in patients with DCI point towards the direction of hypercoagulability compared to patients without DCI (Figure S1; medians (IQR) in Supplemental Material, Table S1, page 1). EXTEM α-angle on admission was significantly higher in patients with DCI than in patients without DCI (p=0.03), but all other parameters, including FIBTEM- and INTEM-parameters, did not differ between the groups at any time point (Supplemental Material, Table S1, page 1). Of conventional coagulation markers, PT (p=0.04) and aPTT (p=0.03) were significantly shorter in DCI patients on admission compared to patients without DCI. Also, D-dimer was significantly higher in DCI patients on days 9-11 (p=0.03) compared to patients without DCI. After exclusion of patients who received tranexamic acid (n=4 patients with DCI and n=22 patients without DCI), the sensitivity analyses showed persistent hypercoagulable ROTEM-profiles on admission and days 3-5 in patients with DCI compared to patients without DCI. Also, EXTEM α-angle on admission remained significantly different (data not shown). On days 9-11, the hypercoagulability shown by some ROTEM-parameters had resolved.

In univariate logistic regression, none of the ROTEM-parameters and conventional coagulation markers were significantly associated with DCI.

### Radiological DCI

Hypercoagulability was observed in patients with radiological DCI, compared to those without (Figure S2; medians (IQR) in Supplemental Material, Table S2, page 2). INTEM CT on admission was significantly shorter compared to those without radiological DCI. On days 3-5, also EXTEM CFT (p=0.03), EXTEM α-angle (p=0.04), EXTEM MCF (p=0.05) and FIBTEM α-angle (p=0.02), FIBTEM A10 (p=0.04) and FIBTEM MCF (p=0.02); and FIBTEM A10 on days 9-11 (p=0.03) were all significantly different in patients with and without radiological DCI. Of conventional coagulation markers, PT (p<0.001) and aPTT (p=0.007) on admission and PT (p=0.02) and fibrinogen (p=0.02) on days 3-5 were significantly different (Figure S2; medians (IQR) in Supplemental Material, Table S2, page 2). In the sensitivity analyses, in which patients who received TXA treatment were excluded (radiological DCI: n=13, no radiological DCI: n=121), all ROTEM-parameters, except from some CT and LI60 parameters, showed consistent hypercoagulability on admission and days 3-5 in patients with radiological DCI compared to patients without. Significance for ROTEM-parameters on days 3-5 was lost. Sensitivity analyses were not done for ROTEM-parameters on days 9-11 as the radiological DCI group included only three patients.

The results of the univariate logistic regression and ROC-curve analyses are listed in Table 2 (ROC-curves in Supplemental Material, Figure S4, page 5). The ROTEM-parameter with the highest AUC for radiological DCI was INTEM CT on admission (AUC: 0.75, 95% C.I. 0.64-0.86), of which the optimal cut-off value was < 153 seconds (sensitivity 94%, specificity 59%).

**Table 2:** Odds ratios, area-under-the-curves and optimal cut-off values with predictive test characteristics of ROTEM-parameters on the occurrence of radiological DCI.

| Variable | OR (95% C.I.) | AUC (95% C.I.) |
| --- | --- | --- |
| **<u>ROTEM-parameters</u>** |  |  |
| T0 INTEM CT | **0.96 (0.94-0.99)** | 0.75 (0.64-0.86) |
| T1 EXTEM CFT | **0.94 (0.89-0.99)** | 0.69 (0.56-0.82) |
| T1 EXTEM α-angle | 1.43 (0.99-2.06) | - |
| T1 EXTEM MCF | 1.16 (1.00-1.34) | - |
| T1 FIBTEM α-angle | 1.25 (0.99-1.57) | - |
| T1 FIBTEM A10 | **1.12 (1.01-1.24)** | 0.71 (0.58-0.83) |
| T1 FIBTEM MCF | **1.12 (1.02-1.24)** | 0.71 (0.58-0.84) |
| **<u>Conventional coagulation markers</u>** |  |  |
| T0 PT | **0.15 (0.04-0.58)** | 0.75 (0.64-0.86) |
| T0 aPTT | **0.68 (0.49-0.93)** | 0.71 (0.57-0.84) |
| T1 PT | **0.18 (0.04-0.77)** | 0.70 (0.54-0.86) |
| T1 APTT | **0.68 (0.47-0.99)** | 0.70 (0.56-0.84) |
| T1 Fibrinogen | **1.55 (1.07-2.26)** | 0.70 (0.55-0.86) |

### Poor clinical outcome

ROTEM-profiles of patients with poor clinical outcome at six months were more hypercoagulable than in patients with good outcome, with increasing differences over time (Figure S3; Supplemental Material, Table A.3, page 3). Patients with poor outcome had significantly shorter EXTEM CT (p=0.01) on admission when compared to patients with good outcome. On days 3-5 and 9-11 all assessed ROTEM-parameters, except from EXTEM CT, EXTEM LI60, INTEM CT and INTEM LI60, showed significant hypercoagulability in patients with poor outcome compared to good outcome. Of conventional coagulation markers, D-dimer on admission (p<0.001); PT (p=0.03), fibrinogen (p=<0.001) and D-dimer (p=0.005) on days 3-5; and aPTT (p=0.004), fibrinogen (p=0.004) and D-dimer (p<0.001) on days 9-11 were significantly different (Figure S3; Supplemental Material, Table A.3, page 3). The sensitivity analyses, in which patients who received TXA treatment were excluded (poor outcome: n=60, good outcome: n=68), showed similar results with sustained statistical significance of the ROTEM-parameters which were significantly different in patients with good and poor clinical outcome in the main analyses (data not shown).

The results of the univariate logistic regression and ROC-curve analyses are listed in Table 3 (ROC-curves in Supplemental Material, Figure S5, page 6-7). The ROTEM-parameters with highest AUC were FIBTEM A10 and FIBTEM MCF on day 3-5 (AUC of both parameters: 0.85, 95% C.I. 0.78-0.92). The optimal cut-off value of FIBTEM A10 was set at > 27 mm (specificity 94%, sensitivity 49%). The optimal cut-off value of FIBTEM MCF was set at > 30 mm (specificity 91%, sensitivity 46%).

**Table 3:** (Adjusted) odds ratios, area-under-the-curves and optimal cut-off values with predictive test characteristics of ROTEM-parameters on poor clinical outcome.

| Variable | OR (95% C.I.) | AUC (95% C.I.) |
| --- | --- | --- |
| **<u>ROTEM-parameters</u>** |  |  |
| T0 EXTEM CT | 1.02 (0.99-1.04) | - |
| T1 EXTEM CFT | **0.91 (0.87-0.95)** | 0.79(0.71-0.88) |
| T1 EXTEM α-angle | **1.72 (1.32-2.24)** | 0.76 (0.66-0.85) |
| T1 EXTEM A10 | **1.27 (1.15-1.41)** | 0.79 (0.71-0.88) |
| T1 EXTEM MCF | **1.36 (1.20-1.54)** | 0.80 (0.72-0.88) |
| T1 INTEM CFT | **0.94 (0.91-0.97)** | 0.76 (0.67-0.86) |
| T1 INTEM α-angle | **1.47 (1.23-1.77)** | 0.76 (0.66-0.85) |
| T1 INTEM A10 | **1.21 (1.12-1.32)** | 0.77 (0.68-0.86) |
| T1 INTEM MCF | **1.30 (1.16-1.45)** | 0.79 (0.71-0.88) |
| T1 FIBTEM α-angle | **1.37 (1.17-1.61)** | 0.76 (0.67-0.85) |
| T1 FIBTEM A10 | **1.31 (1.18-1.45)** | 0.85 (0.78-0.92) |
| T1 FIBTEM MCF | **1.28 (1.17-1.40)** | 0.85 (0.78-0.92) |
| T2 EXTEM CFT | **0.93 (0.86-0.99)** | 0.74 (0.60-0.87) |
| T2 EXTEM α-angle | **1.46 (1.02-2.09)** | 0.71 (0.57-0.85) |
| T2 EXTEM A10 | **1.18 (1.03-1.34)** | 0.74 (0.61-0.88) |
| T2 EXTEM MCF | **1.23 (1.04-1.45)** | 0.71 (0.57-0.85) |
| T2 INTEM CFT | **0.95 (0.91-0.998)** | 0.68 (0.54-0.83) |
| T2 INTEM α-angle | **1.35 (1.04-1.77)** | 0.68 (0.54-0.82) |
| T2 INTEM A10 | **1.14 (1.02-1.28)** | 0.72 (0.58-0.87) |
| T2 INTEM MCF | **1.19 (1.03-1.38)** | 0.71 (0.57-0.85) |
| T2 FIBTEM α-angle | **1.39 (1.08-1.79)** | 0.72 (0.59-0.86) |
| T2 FIBTEM A10 | **1.27 (1.11-1.46)** | 0.82 (0.72-0.93) |
| T2 FIBTEM MCF | **1.26 (1.10-1.43)** | 0.82 (0.72-0.93) |
| **<u>Conventional coagulation markers</u>** |  |  |
| T0 D-dimer | **1.11 (1.04-1.18)** | 0.69 (0.60-0.78) |
| T1 PT | **0.45 (0.21-0.94)** | 0.63 (0.52-0.73) |
| T1 Fibrinogen | **2.43 (1.74-3.40)** | 0.84 (0.77-0.92) |
| T1 D-dimer | 1.26 (0.92-1.74) | - |
| T2 APTT | 0.62 (0.43-0.90) | - |
| T2 Fibrinogen | **1.87 (1.18-2.96)** | 0.73 (0.60-0.86) |
| T2 D-dimer | 1.02 (0.99-1.04) | - |

## DISCUSSION

In this study, ROTEM-profiles of aSAH patients with DCI, radiological DCI and poor outcome at follow-up were hypercoagulable when compared to patients without DCI, radiological DCI and poor outcome, throughout the entire time course after SAH. For radiological DCI, INTEM CT on admission and FIBTEM α-angle, A10 and MCF on days 3-5 had an acceptable discriminative ability (AUC > 0.70). For poor outcome, nearly all markers of clot kinetics and clot strength) of assays at days 3-5 and days 9-11 had an AUC > 0.70. The ROTEM-parameter with best discriminative ability for radiological DCI was INTEM CT on admission < 153 seconds, and for poor outcome FIBTEM A10 at days 3-5 after aSAH with an optimal cut-off value of > 27 mm. Of note, these cut-off values are within the range of what is considered ‘normal’. However, the episode following aSAH cannot be considered a ‘normal’ episode and this study provides novel insights into potentially unsafe ranges following aSAH.

### DCI

All but one ROTEM-parameters showed relative hypercoagulability in patients with DCI compared to those without DCI, which was even more pronounced in patients who had radiological DCI compared to those without. The more pronounced hypercoagulability found in radiological DCI may be explained by the use of objective diagnostic criteria and emphasizes the shortcomings of the current clinical diagnosis-per-exclusionem of DCI, which likely has led to a heterogenic group of patients who might have had complications other than cerebral ischemia. The ROTEM-parameter with best discriminative power was INTEM CT on admission. Interestingly, on day 3-5 hypercoagulability was observed by multiple EXTEM- and FIBTEM-parameters on day 3-5, which suggests an increased contribution of fibrinogen and platelet activation to clot formation. This is in line with the results of the two previous studies on ROTEM in DCI, which showed significant hypercoagulability in the FIBTEM assay, although the sample timing differed somewhat from our study.^18,20^ After exclusion of patients who received tranexamic acid, the deranged ROTEM-parameters on days 3-5 and 9-11 lost statistical significance. This may be explained by loss of power resulting from smaller sample sizes. Potential confounding by tranexamic acid is less likely, as the proportion of patients treated with tranexamic acid in patients without DCI was larger than in patients with DCI. Also, ROTEM parameters of lysis did not differ between groups.

In current clinical practice, all SAH patients are hospitalized for a minimum of two weeks for monitoring for DCI. ROTEM may aid in the decision whom to safely discharge at an earlier time. The determination of an optimal cut-off point with a high sensitivity is somewhat offset by poor specificity. However, in case of considering earlier discharge, poor specificity may be of little relevance, as these patients would otherwise have been admitted for a longer duration anyway. The ROTEM-parameter with highest discriminative ability of excluding DCI was observed for INTEM CT on admission, with a cut-off point of < 153 seconds. Potentially, a combination of ROTEM-parameters with clinical characteristics or other laboratory markers will further improve the discriminative ability.

Conventional coagulation markers PT and aPTT on admission and on days 3-5, were significantly shorter with acceptable discriminative ability to detect patients with radiological DCI. Also, fibrinogen on days 3-5 was significant higher in patients with than without radiological DCI. However, later in the course of SAH, these differences disappeared, whereas differences in FIBTEM-parameters became more outspoken. Previous studies on associations between DCI and PT or aPTT have shown inconsistent results, which hampers the role for PT and aPTT in the prediction of DCI.^12^

### Poor outcome

Hypercoagulable profiles as assessed by ROTEM have a high discriminative ability to prognosticate poor outcome. The hypercoagulable profiles were determined by parameters of clot kinetics and clot strength on days 3-5 and 9-11, indicating increased platelet activity and fibrinogen contribution to blood clot formation, which is in line with previous studies using rotational thromboelastography (TEG, a comparable viscoelastic test).^14,28–31^ FIBTEM markers yielded the highest area-under-the-curves, with comparable discriminative ability to the most commonly used prognosticator, the WFNS grade.^32,33^ The contribution of fibrinogen to poor clinical outcome in aSAH may be in line with recent insights into the role of fibrinogen in amplifying immune and degenerative processes in the brain in various neurological diseases and following neurotrauma.^34^ Whether correction of the observed hypercoagulability will result in improved clinical outcome is yet unknown. Based on our results, it is tempting to speculate that anti-platelet drugs, fibrinogen-depleting drugs or inhibitors of fibrin formation might be potential treatment strategies to reduce poor clinical outcome after aSAH. In the past, a Cochrane review on anti-platelet therapy after SAH showed a non-significant trend towards a lower occurrence of DCI and improved clinical outcome, but was possibly offset by a concomitant non-significant increase in haemorrhagic complications.^35^ ROTEM might therefore be helpful in stratifying patients with hypercoagulability for therapeutic interventions, without exposing patients without hypercoagulability to an increased risk of haemorrhagic complications.

Naturally, our study has some limitations. The number of patients with DCI, and especially radiological DCI, is limited. The cut-off values were determined using a sensitivity threshold of at least 90%. In order to discharge patients at low risk of DCI, or to guide physicians to determine the futility of care, an even higher sensitivity with preservation of specificity would be preferable. In our relatively small cohort this was not possible, however larger cohorts will allow for the determination of more optimal cut-off points. Strengths are the prospective nature of our study, the use of standardized outcome measurements, and the evaluation of longitudinally measured ROTEM-parameters during the first two weeks following aSAH. To the best of our knowledge, this is the first study to assess the discriminative ability of ROTEM-parameters for DCI and poor clinical outcome.

Hypercoagulability, as detected by ROTEM-parameters, is an excellent marker of poor clinical outcome. In combination with other clinical and laboratory parameters, ROTEM-parameters might aid physicians in their decisions to forego life-sustaining treatments. Also, ROTEM-parameters could potentially be useful in stratifying aSAH patients with hypercoagulability for future trials on therapeutic interventions with the aim to improve clinical outcome. Also, ROTEM-parameters might be useful in stratifying patients with low risk of developing radiological DCI for earlier discharge.

## Authorship details

**M.A. Tjerkstra:** concept and design, analysis and/or interpretation of data, critical writing the intellectual content, final approval of the version; **H. Labib, B. A. Coert and R. Post**: revising the intellectual content, final approval of the version; **W. P. Vandertop, D. Verbaan and N.P. Juffermans:** concept and design, analysis and/or interpretation of the data, revising the intellectual content, final approval of the version.

## Data Availability

Data are available on request, with the request submitted to the corresponding author for consideration.

## References

1. D’Souza S. Aneurysmal Subarachnoid Hemorrhage. J Neurosurg Anesthesiol. 2015;27:222-240. doi: 10.1097/ana.0000000000000130

2. Stienen MN, Germans M, Burkhardt J-K, Neidert MC, Fung C, Bervini D, Zumofen D, Roethlisberger M, Marbacher S, Maduri R, et al. Predictors of In-Hospital Death After Aneurysmal Subarachnoid Hemorrhage. Stroke. 2018;49:333–340. doi: doi:10.1161/STROKEAHA.117.019328

3. Kassell NF, Torner JC, Haley EC, Jr., Jane JA, Adams HP, Kongable GL. The International Cooperative Study on the Timing of Aneurysm Surgery. Part 1: Overall management results. Journal of neurosurgery. 1990;73:18-36. doi: 10.3171/jns.1990.73.1.0018

4. Hop JW, Rinkel GJ, Algra A, van Gijn J. Case-fatality rates and functional outcome after subarachnoid hemorrhage: a systematic review. Stroke. 1997;28:660–664.

5. Roos Y, De Haan R, Beenen L, Groen R, Albrecht K, Vermeulen M. Complications and outcome in patients with aneurysmal subarachnoid haemorrhage: a prospective hospital based cohort study in the Netherlands. J Neurol Neurosurg Psychiatry. 2000;68:337–341.

6. Budohoski KP, Guilfoyle M, Helmy A, Huuskonen T, Czosnyka M, Kirollos R, Menon DK, Pickard JD, Kirkpatrick PJ. The pathophysiology and treatment of delayed cerebral ischaemia following subarachnoid haemorrhage. Journal of neurology, neurosurgery, and psychiatry. 2014:jnnp-2014-307711.

7. Vergouwen MD, Vermeulen M, Coert BA, Stroes ES, Roos YB. Microthrombosis after aneurysmal subarachnoid hemorrhage: an additional explanation for delayed cerebral ischemia. J Cereb Blood Flow Metab. 2008;28:1761–1770.

8. Geraghty JR, Testai FD. Delayed Cerebral Ischemia after Subarachnoid Hemorrhage: Beyond Vasospasm and Towards a Multifactorial Pathophysiology. Current Atherosclerosis Reports. 2017;19:50. doi: 10.1007/s11883-017-0690-x

9. Sabri M, Ai J, Lakovic K, D’abbondanza J, Ilodigwe D, Macdonald R. Mechanisms of microthrombi formation after experimental subarachnoid hemorrhage. Neuroscience. 2012;224:26–37.

10. Connolly ES, Rabinstein AA, Carhuapoma JR, Derdeyn CP, Dion J, Higashida RT, Hoh BL, Kirkness CJ, Naidech AM, Ogilvy CS, et al. Guidelines for the Management of Aneurysmal Subarachnoid Hemorrhage A Guideline for Healthcare Professionals From the American Heart Association/American Stroke Association. Stroke. 2012;43:1711–1737. doi: 10.1161/STR.0b013e3182587839

11. Steiner T, Juvela S, Unterberg A, Jung C, Forsting M, Rinkel G. European Stroke Organization Guidelines for the Management of Intracranial Aneurysms and Subarachnoid Haemorrhage. Cerebrovasc Dis. 2013;35:93–112. doi: 10.1159/000346087

12. Boluijt J, Meijers JC, Rinkel GJ, Vergouwen MD. Hemostasis and fibrinolysis in delayed cerebral ischemia after aneurysmal subarachnoid hemorrhage: a systematic review. J Cereb Blood Flow Metab. 2015;35:724–733.

13. Luddington RJ. Thrombelastography/thromboelastometry. Clin Lab Haematol. 2005;27:81-90. doi: 10.1111/j.1365-2257.2005.00681.x

14. Tjerkstra MA, Wolfs AE, Verbaan D, Vandertop WP, Horn J, Müller MCA, Juffermans NP. A systematic review on viscoelastic testing in subarachnoid haemorrhage patients. World Neurosurg. 2023. doi: 10.1016/j.wneu.2023.03.108

15. Baranich AI, Polupan AA, Sychev AA, Savin IA. Thromboelastometry as a Comprehensive Assessment of Hypercoagulation After Aneurysmal Subarachnoid Hemorrhage: A Case Report and Literature Review. Subarachnoid…. 2019.

16. Builes JCG, Reverter PT. Coagulation profile evaluated by thromboelastography rotem® in patients with subarachnoid hemorrhage admitted to an intensive care unit. Intensive care…. 2015.

17. Hvas CL, Lauridsen SV, Pedersen ES, Gyldenholm T, Hvas AM. Ex vivo effect of hemostatic therapy in subarachnoid and intracerebral hemorrhage. Thromb Res. 2020;189:42–47. doi: 10.1016/j.thromres.2020.02.012

18. Lauridsen SV, Hvas CL, Sandgaard E, Gyldenholm T, Mikkelsen R, Obbekjaer T, Sunde N, Tonnesen EK, Hvas AM. Thromboelastometry Shows Early Hypercoagulation in Patients with Spontaneous Subarachnoid Hemorrhage. World Neurosurg. 2019;130:e140–e149. doi: 10.1016/j.wneu.2019.06.019

19. Ettinger MG. Coagulation abnormalities in subarachnoid hemorrhage. Stroke. 1970;1:139–142.

20. Vahtera AS, Junttila EK, Jalkanen LV, Huhtala HS, Katanandova KV, Helen PT, Kuitunen AH. Activation of Blood Coagulation After Aneurysmal Subarachnoid Hemorrhage: A Prospective Observational Trial of Rotational Thromboelastometry. World Neurosurg. 2019;122:e334–e341. doi: 10.1016/j.wneu.2018.10.035

21. Post R, Germans MR, Tjerkstra MA, Vergouwen MDI, Jellema K, Koot RW, Kruyt ND, Willems PWA, Wolfs JFC, de Beer FC, et al. Ultra-early tranexamic acid after subarachnoid haemorrhage (ULTRA): a randomised controlled trial. The Lancet. 2021;397:112–118. doi: 10.1016/S0140-6736(20)32518-6

22. Vergouwen MD, Vermeulen M, van Gijn J, Rinkel GJ, Wijdicks EF, Muizelaar JP, Mendelow AD, Juvela S, Yonas H, Terbrugge KG. Definition of delayed cerebral ischemia after aneurysmal subarachnoid hemorrhage as an outcome event in clinical trials and observational studies: proposal of a multidisciplinary research group. Stroke. 2010;41:2391–2395.

23. Hosmer Jr DW, Lemeshow S, Sturdivant RX. Applied logistic regression. John Wiley & Sons; 2013.

24. Deeks JJ, Altman DG. Diagnostic tests 4: likelihood ratios. BMJ. 2004;329:168–169. doi: 10.1136/bmj.329.7458.168

25. Ranganathan P, Aggarwal R. Common pitfalls in statistical analysis: Understanding the properties of diagnostic tests - Part 1. Perspect Clin Res. 2018;9:40–43. doi: 10.4103/picr.PICR_170_17

26. Ranganathan P, Aggarwal R. Understanding the properties of diagnostic tests - Part 2: Likelihood ratios. Perspect Clin Res. 2018;9:99–102. doi: 10.4103/picr.PICR_41_18

27. Trevethan R. Sensitivity, Specificity, and Predictive Values: Foundations, Pliabilities, and Pitfalls in Research and Practice. Frontiers in Public Health. 2017;5. doi: 10.3389/fpubh.2017.00307

28. Espino E, Torres GL, Savarraj J, Bajgur S, Choi HA. Thromboelastography following spontaneous subarachnoid hemorrhage. Neurocrit Care. 2015;23:S209. doi: 10.1007/s12028-015-0193-y

29. Frontera JA, Provencio JJ, Sehba FA, McIntyre TM, Nowacki AS, Gordon E, Weimer JM, Aledort L. The Role of Platelet Activation and Inflammation in Early Brain Injury Following Subarachnoid Hemorrhage. Neurocrit Care. 2017;26:48–57. doi: 10.1007/s12028-016-0292-4

30. Ramchand P, Nyirjesy S, Frangos S, Doerfler S, Nawalinski K, Quattrone F, Ju C, Patel H, Driscoll N, Maloney-Wilensky E, et al. Thromboelastography Parameter Predicts Outcome After Subarachnoid Hemorrhage: An Exploratory Analysis. World Neurosurg. 2016;96:215–221. doi: 10.1016/j.wneu.2016.04.002

31. Zeineddine HA, Li W, T PK, McBride D, Dienel A, Torres G, Grotta J, Savarraj J, Chang T, Choi HA, et al. Thromboelastography Indices for Predicting Outcomes After Aneurysmal Subarachnoid Hemorrhage: A Prospective Study. Stroke. 2022;53:e221–e223. doi:10.1161/STROKEAHA.122.039372

32. Nguyen TA, Vu LD, Mai TD, Dao CX, Ngo HM, Hoang HB, Do SN, Nguyen HT, Pham DT, Nguyen MH, et al. Predictive validity of the prognosis on admission aneurysmal subarachnoid haemorrhage scale for the outcome of patients with aneurysmal subarachnoid haemorrhage. Sci Rep. 2023;13:6721. doi: 10.1038/s41598-023-33798-5

33. van Heuven AW, Dorhout Mees SM, Algra A, Rinkel GJ. Validation of a prognostic subarachnoid hemorrhage grading scale derived directly from the Glasgow Coma Scale. Stroke. 2008;39:1347–1348. doi: 10.1161/strokeaha.107.498345

34. Petersen MA, Ryu JK, Akassoglou K. Fibrinogen in neurological diseases: mechanisms, imaging and therapeutics. Nature Reviews Neuroscience. 2018;19:283–301. doi: 10.1038/nrn.2018.13

35. Dorhout Mees SM, Van Den Bergh WM, Algra A, Rinkel GJE. Antiplatelet therapy for aneurysmal subarachnoid haemorrhage. Cochrane Database of Systematic Reviews. 2007;(2) (no pagination). doi: 10.1002/14651858.CD006184.pub2

